## Supplementary Information for "Combining participatory mapping and route optimization algorithms to inform the delivery of community health interventions at the last mile"

**Additional file 1: Workflow for VRPTW algorithm implementation**

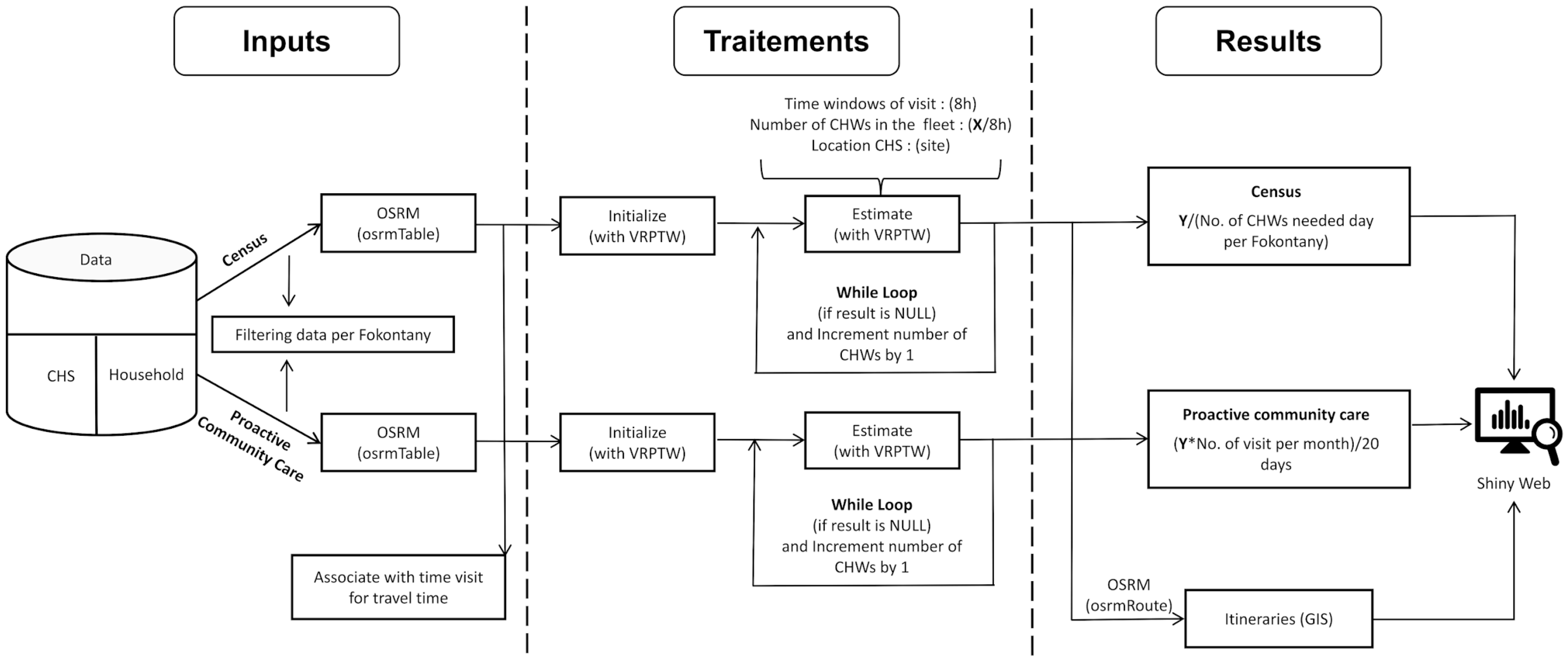

- Data Geograprhics
- Data «Time Matrix» for Time Travel Time
- Data Time Visit

- **Initialize**, Travel Time of All Routes for one Person, Without Time Constraints (Result Variable **X**).
- **Estimate**, Travel Time of All Routes Defined Per Person, With a Time Constraint of 8 Hours (Result Variable **Y**).

- Identification Person-Day, Month per Fokontany
- Road CHW to Visit Houshold
- Vizualisation of Results and Proactive Community Care Planning Tools.

**Additional file 2: Estimated number of personnel required per commune for mass distribution campaigns.**

| Commune | Number of buildings | Travel distance (km) | Travel duration (h) | Personnel -Day | Personnel -Month |
|---|---|---|---|---|---|
| Ambiabe | 3,583 | 1,767 | 1,222 | 163 | 11 |
| Ambohimanga Du Sud | 13,050 | 4,107 | 3,987 | 533 | 37 |
| Ambohimiera | 11,511 | 4,759 | 3,746 | 501 | 34 |
| Ampasinambo | 2,975 | 487 | 400 | 56 | 5 |
| Analampasina | 5,199 | 2,186 | 1,700 | 227 | 14 |
| Androrangavola | 8,519 | 3,433 | 2,756 | 368 | 27 |
| Antaretra | 4,636 | 1,524 | 1,430 | 193 | 15 |
| Antsindra | 5,422 | 2,669 | 1,851 | 245 | 15 |
| Fasintsara | 5,136 | 1,640 | 1,574 | 218 | 19 |
| Ifanadiana | 7,478 | 3,796 | 2,574 | 339 | 20 |
| Kelilalina | 5,347 | 2,000 | 1,697 | 229 | 18 |
| Maroharatra | 9,032 | 3,582 | 2,906 | 393 | 26 |
| Marotoko | 5,370 | 2,244 | 1,753 | 235 | 17 |
| Ranomafana | 4,182 | 1,797 | 1,372 | 184 | 13 |
| Tsaratanana | 16,488 | 8,505 | 5,706 | 755 | 48 |
| **Total** | **107,928** | **44,496** | **34,674** | **4,639** | **319** |

**Additional file 3 : Estimated number of personnel required per commune for proCCM programs.**

| | | Visit - once per month | | Visit - twice per month | | Visit - four times per month | |
|---|---|---|---|---|---|---|---|
| Commune | Number of households | Personnel | | Personnel | | Personnel | |
| | | Day (Ref) | Month | Day | Month | Day | Month |
| Ambiabe | 1,433 | 55 | 7 | 110 | 8 | 220 | 12 |
| Ambohimanga Du Sud | 5,221 | 171 | 23 | 342 | 27 | 684 | 45 |
| Ambohimiera | 4,604 | 163 | 21 | 326 | 27 | 652 | 41 |
| Ampasinambo | 496 | 19 | 5 | 38 | 5 | 76 | 5 |
| Analampasina | 2,081 | 73 | 9 | 146 | 11 | 292 | 18 |
| Androrangavola | 3,406 | 119 | 15 | 238 | 18 | 476 | 31 |
| Antaretra | 1,855 | 60 | 10 | 120 | 10 | 240 | 15 |
| Antsindra | 2,168 | 81 | 7 | 162 | 12 | 324 | 20 |
| Fasintsara | 2,054 | 77 | 16 | 154 | 17 | 308 | 22 |
| Ifanadiana | 2,993 | 112 | 10 | 224 | 16 | 448 | 27 |
| Kelilalina | 2,138 | 72 | 12 | 144 | 14 | 288 | 20 |
| Maroharatra | 3,616 | 129 | 19 | 258 | 23 | 516 | 34 |
| Marotoko | 2,147 | 76 | 10 | 152 | 12 | 304 | 18 |
| Ranomafana | 1,673 | 60 | 8 | 120 | 10 | 240 | 14 |
| Tsaratanana | 6,596 | 241 | 24 | 482 | 34 | 964 | 57 |
| **Total** | **42,481** | **1,508** | **196** | **3,016** | **244** | **6,032** | **379** |

**Additional file 4: Correlation between daily personnel requirements and CHW catchment characteristics in Ifanadiana district.**

A-C: Mass administration campaigns, D-F: Proactive Community Health (1 visit per month). Panels A and D plot a linear regression line due to the nature of the relationship, while the other panels use non-linear local regression fitting.

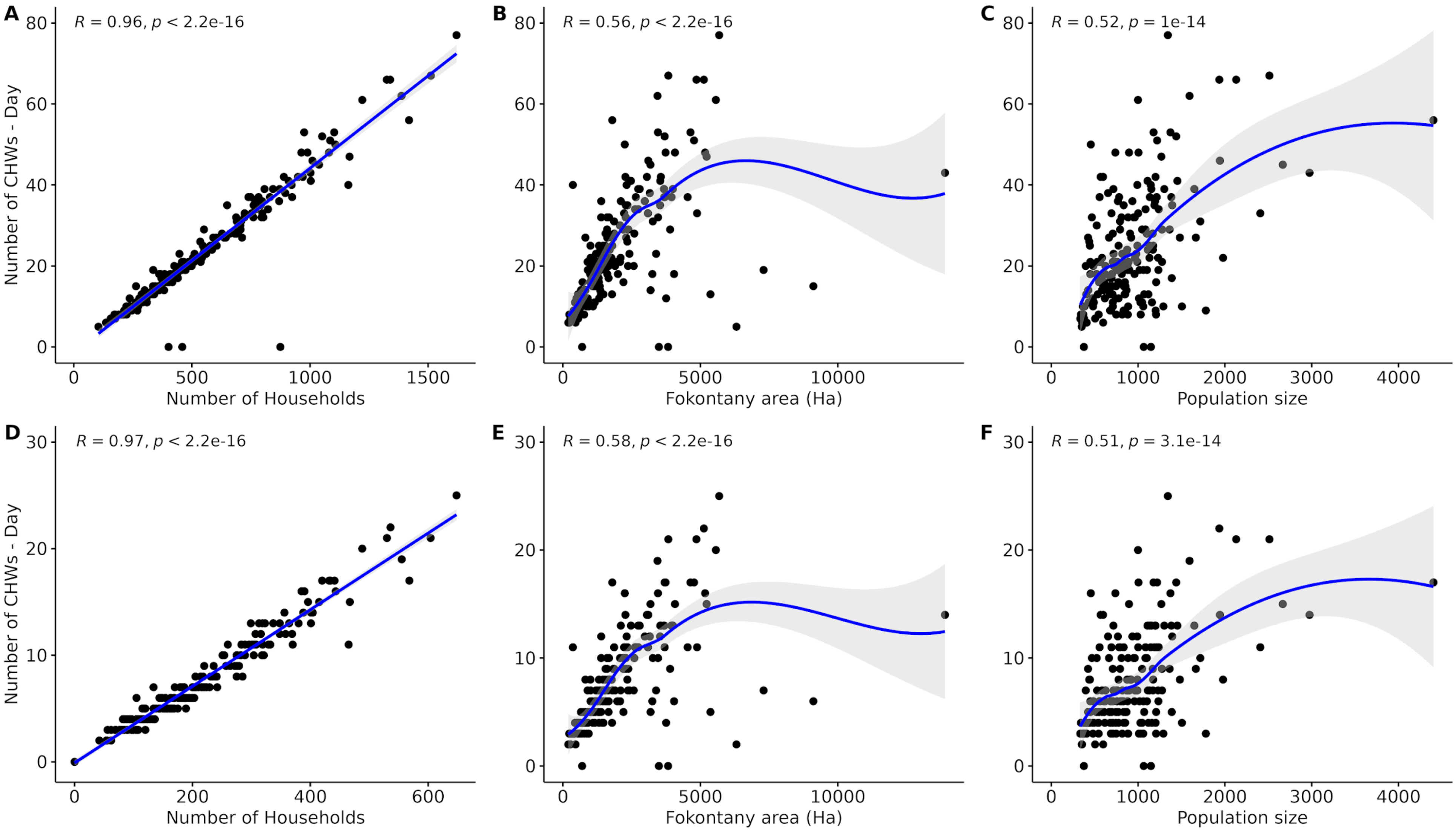

### Additional file 5: Comparison of algorithm predictions and field-observed CHW needs in Ifanadiana district

Predictions obtained from the VRPTW algorithm were compared to results obtained in the field in terms of personnel-days required to understand the validity of our hypotheses and the validity of the results generated. For this, we took advantage of a previous census survey that was conducted in two communes of Ifanadiana District (Kelilalina and Tsaratanana) back in 2020 by teams from the NGO Pivot and CHWs from those communes. Since these surveys were conducted two years prior to the development of our algorithms, data collection had not been specifically recorded with the level of detail necessary for a robust validation of the tool. Instead, a group session was organized to facilitate discussions between the authors and 14 participants from the community health teams who had participated in the field surveys. The objective of this group session was to gather information regarding the human resources deployed per commune, as well as the number of days of work and the average duration of work that had been necessary for carrying out these census surveys.

The census was achieved with a team of 35 personnel working over a period of 6 days for about 11 hours per day in the Kelilalina commune, and a team of 47 personnel working over a period of 18 days for about 10 hours per day in the Tsaratanana commune. Subsequently, we standardized the calculations in terms of personnel-days to make them comparable with our own calculations (assuming a worker puts in 8 hours of work per day), which resulted in 289 personnel-days for Kelilalina and 1,058 personnel-days for Tsaratanana. To compare these observed values with the predictions obtained from the VRPTW algorithm, we used the parameters of the mass distribution campaign scenario for these two communes, which is equivalent to a full census in that all buildings in the catchment are visited and those who are inhabited households require a higher visit time.

**Table. Comparison of algorithm predictions and field-observed CHW needs in Ifanadiana district**

| Commune | Number of buildings | Personnel-Days | |
|---|---|---|---|
| | | Observed | VRPTW Prediction (% difference prediction vs. observed) |
| Kelilalina | 5,347 | 289 | 229 (-21%) |
| Tsaratanana | 16,488 | 1,058 | 755 (-29%) |

When VRPTW predictions were compared with observed personnel-days during field surveys, the optimization algorithm predicted a 29% and 21% decrease in the number of resources necessary in Tsaratanana and Kelilalina, respectively. This result was in line with our expectations, given that CHW itineraries had not been optimized for geography during fieldwork, and highlights potential benefits of the optimization algorithm for planning and scheduling.
